# Impact of a one-year interruption to vector control on Bioko Island, Equatorial Guinea

**DOI:** 10.1101/2025.11.04.25339542

**Authors:** David S. Galick, Dianna E.B. Hergott, Norberto Bosepa Cubacuba, Teresa Ayingono Ondo Mifumu, Jordan M. Smith, Matilde Riloha Rivas, Wonder P. Phiri, David L. Smith, Carlos A. Guerra, Guillermo A. García

## Abstract

**Background:** Over the last 20 years, malaria transmission on Bioko Island, Equatorial Guinea has declined dramatically thanks to the implementation of a comprehensive set of malaria control activities, centered around island-wide indoor residual spraying (IRS). In 2024 Bioko Island experienced a lapse in malaria control funding, and as a result vector control activities (including IRS) were interrupted. However, agreements with funders allowed for both a previously planned malaria indicator survey (MIS) and the subsequent implementation of indoor residual spraying (IRS) in late 2024.

**Methods:** This study analyses routine case data from public health facilities from 2019-2024 and annual cross-sectional MIS data from 2019-2024 using interrupted time series methods to quantify the impact of the interruption and reestablishment of malaria control activities on Bioko Island. Modeling included interaction terms to allow both pre-intervention trends and interruption effect sizes to vary by district.

**Results:** In 2024, the number of confirmed cases reported was 41% higher than the 2021-2023 average, and the *Plasmodium falciparum* prevalence rate (p*f*PR) rose by three percentage points. Statistical modeling estimated that 23.1% (95% CI 9.2-33.7%) of 2024 cases were associated with the timing of vector control interruptions and potentially avertable if control activities had been maintained, and that the interruption was associated with an increased P*f*PR in 2024, above previous trends (adjusted OR 1.20, 95% CI 1.07-1.35). Moreover, the reintroduction in IRS in late 2024 was found to have potentially averted an additional 13.3% (95% CI 4.4-28.4%) increase in confirmed cases.

**Conclusions:** In just one year with interruptions to control, malaria transmission and burden quickly resurged on Bioko Island. However, Bioko’s experience demonstrates that the reestablishment of control activities can equally rapidly contain, and reverse resurgence associated with control interruptions.

## Introduction

Since 2000, a renewed commitment to malaria control has substantially reduced the number of cases and deaths caused by malaria worldwide, and especially in sub-Saharan Africa [1]. These gains were achieved primarily through intensive, widespread vector control activities, including both the distribution of long-lasting insecticide-treated nets (LLINs) and administration of indoor residual spraying (IRS) [2]. However, since 2015, progress has stalled, in part due to funding gaps, challenges to the effectiveness of existing tools, and a lack of new, scalable tools currently available for programmatic use [3,4]. These challenges serve as a reminder of the vulnerability of progress already made, especially in the face of interruptions to control activities.

The impacts of malaria control interruptions have long been clear. Historically, funding disruptions have been the most common cause of malaria resurgence across world regions [5]. In recent years, re-allocation of funding and external shocks such as the COVID-19 pandemic have demonstrated again the fragility of reductions in malaria transmission and burden to even partial disruptions of control activities [6–8]. Multiple modeling exercises have also explored and estimated the enormous additional burden that large interruptions in malaria control activities could cause [9–11].

Despite the overwhelming evidence that funding reductions on malaria control in areas with ongoing transmission will result in significant resurgence, the funding environment remains difficult due to political, economic, and other concerns. Given this situation, it is crucial to measure and document the effects of funding reductions when they occur, where possible. The results from such a funding disruption on Bioko Island, Equatorial Guinea in 2024 are reported here. Thanks to the continued reporting of routine data in the National Health Information System, and re-establishment of funding in late 2024 in time to conduct a malaria indicator survey (MIS) as previously planned, quantification of the impact of the interruption using interrupted time-series methods on both malaria cases and prevalence was possible.

## Methods

### Study area

Bioko Island is the largest island of Equatorial Guinea, located in the Bight of Biafra off the coast of Cameroon. The island’s population is approximately 270,000, predominantly concentrated in the capital city, Malabo [12,13]. Bioko has historically high and perennial malaria transmission, but the implementation of a robust malaria control project based on island-wide indoor residual spraying (IRS) in 2004 dramatically reduced malaria transmission and burden [14,15]. Since 2015, vector control strategies have changed: from 2015-2020 IRS was targeted to only high transmission areas, supported by island-wide long-lasting insecticide treated net (LLIN) mass distribution campaigns in 2015 and 2018, while since 2021 LLIN distribution has been conducted primarily via continuous distribution channels and IRS re-expanded to target the entire island at varied coverage levels [16,17]. In 2022-2023, the vector control portfolio also incorporated increasingly larger scale larval source management (LSM) activities, particularly in the greater Malabo area. These vector control interventions have been supported by case management activities (including providing free diagnosis and treatment of malaria in public health facilities) and social behavioral change communication activities.

In January 2024, because of delays in funding negotiations, there was an interruption of most malaria control activities. All planned field activities, including the implementation of IRS and LSM, and operation of continuous LLIN distribution points were delayed or canceled, regardless of existing stock of materials. Only routine activities with decentralized implementation, such as the diagnosis and treatment of cases, and distribution of LLINs to pregnant women during antenatal care, were able to continue. This interruption continued until July, when an agreement was reached with funders to support the implementation of key malaria prevention activities, including both IRS and LLIN continuous distribution, but not LSM. Funders also agreed to support implementation of the annual malaria indicator survey (MIS) as originally planned, in August-September, in order to better quantify the impact of the interruption. As a result, LLIN distribution points in Malabo district reopened in July, followed by implementation of the MIS beginning in August, while IRS was conducted from late September through December. LLIN distribution points in other districts were opened in October-November, corresponding closely with the implementation of IRS in these districts. Due to a shortage of insecticide on hand, not all areas of the island could be targeted with IRS, so some with historically low transmission and high housing quality were excluded. In all targeted communities, IRS implementation followed the completion of MIS data collection.

### Routine health data

Public health facilities on Bioko report routine indicators, including confirmed malaria cases (confirmed by RDT or microscopy), to the national malaria control program (NMCP), which are subsequently digitized into a DHIS2 system. For data reported prior to October 2023, individual-level records were digitized using a Tracker module, but due to the large workload of data digitization, the NMCP implemented an aggregate data collection module starting in October 2023, which enabled much more rapid data entry while still providing key indicators. All health facilities reported data during every month of active operation during the study period. Here, aggregated monthly all-age monthly reported confirmed malaria cases for each of the four districts on Bioko Island (Malabo, Baney, Luba, and Riaba) from January 2021 to December 2024 were extracted for analysis, and divided by annual district-level population estimates (see below) to compute case incidence per 1,000. These data include reported cases from all public health facilities on Bioko Island active during the corresponding month and did not contain any identifiable information. While malaria-attributable hospitalizations and deaths are also collected, these data were not analyzed due to the relatively low number reported and potential changes in definitions used during the study period. Given the moderate level of on-island malaria transmission, travel histories are not collected as part of routine malaria surveillance.

An important change in the health system on Bioko Island occurred during the study period: a new public hospital, the General Hospital of Sampaka, opened for services in January 2024. During the first six months of 2024, the number of patients attended in Sampaka slowly grew but remained relatively small. In the second half of the year, however, the volume of patients significantly increased after pediatric services were transferred from the Regional Hospital of Malabo (the main public tertiary facility on Bioko) to be provided instead in Sampaka.

### Malaria indicator surveys

Malaria Indicator Surveys conducted on Bioko Island from 2019-2024 were the primary source of prevalence and covariate data in this study. The methodology of the annual Bioko Island MIS has been published elsewhere [18]. In brief, the questionnaire was adapted from the RBM MIS toolkit [19], and a stratified cluster sampling design based on 109 primary sampling units (PSUs) covering all inhabited areas of Bioko was used. Strata were defined according to the population density and residual local transmission as modeled in 2018 [20], and target sample sizes were set at 25% of inhabited households for PSUs in the rural/high transmission stratum, and 5% of inhabited households for PSUs in the urban/low transmission stratum for each year. The sampling frame was updated each year based on a household census conducted during an annual round of IRS (conducted between February and July), and survey interviews were conducted from August to early October each year, following a standardized timeline to improve comparability of estimates. All consenting household members (regardless of age) were tested for malaria using a rapid diagnostic test (STANDARD Q Malaria Pf/Pan RDT, SD BIOSENSOR). Covariates used from the MIS in this analysis included self-reported LLIN use and time of house entry on the night preceding the survey interview, history of travel outside Bioko Island in the 8 weeks preceding the survey interview, and socioeconomic status (SES) based on principal components analysis of household possessions, grouped into quintiles. Note that LLIN use and house entry time questions were asked only for participants who reported sleeping at home the night preceding the survey interview, and this was the main cause of missing data in this study.

### Other data

Precipitation data were derived from a global satellite-sensed precipitation product with 0.1-degree resolution [21]. Daily precipitation estimates were down-sampled to 1km × 1km grid cells (map areas) by bilinear interpolation, and for the cases analysis these estimates were weighted by the number of inhabited houses in each map area according to the 2024 IRS campaign data to generate population-weighted district precipitation estimates. Population estimates used here were based on data from a LLIN mass distribution campaign conducted in 2018. [13] District-level population estimates for 2018 population were extracted and extrapolated to each year in the study period, assuming an annual growth rate of 2.5%. Annual IRS coverage and LSM targeting from 2019-2023 at the PSU-level, and cumulative IRS coverage and cumulative LLINs distributed in continuous distribution points by month and district in 2024 were extracted from campaign data.

### Statistical analysis

Interrupted time series methods were used for the analysis of both routine health system and survey data. Analyses were conducted in a manner to allow for the possibility that the impacts in each district could be distinct. District-level estimates were then aggregated to compute island-wide estimates and confidence intervals.

For analysis of reported confirmed cases, a negative binomial generalized additive model with log link was fit to case data, incorporating low-rank splines (4 degrees of freedom) to represent the long-term (interannual) trend and impact of the interruption of services in 2024. Smoothing parameters were selected automatically [22], but sensitivity analysis on the degrees of freedom was not conducted since increasing would substantially impact model identifiability. Model fitting was performed on the case incidence per 1,000 population scale. Precipitation aggregated to monthly scale centered around the 1^st^ of the month (i.e. aggregating 16 August – 15 September for the value of September), with lags of 0,1 and 2 months were included as covariates. To assess the possible impact of the reestablishment of indoor residual spraying (IRS) in late 2024, cumulative achieved 2024 IRS coverage was also included as a covariate, calculated as the proportion of inhabited houses sprayed by the beginning of each month. This covariate was extracted from IRS campaign data at district level and overall, leveraging the underlying spatial decision support system which has been described in detail elsewhere [23]. The cumulative number of LLINs distributed in continuous distribution points since re-opening in 2024 was also included as a potential covariate in initial modeling but excluded from the final model since models including LLIN distribution performed more poorly according to AIC. Since LSM was not conducted in 2024, the effect of its reintroduction could not be assessed. All effects and the intercept were modeled as interactions with district to allow for variation in pre-interruption trends, the impacts of the interruption and re-establishment of control. Residuals were examined for autocorrelation, and a sensitivity analysis accounting for potential autocorrelation was performed (see Supplementary Information)

Fitted models were used to estimate the incidence under two counterfactual scenarios: 1) that there was no interruption of services in 2024 (CF1), and 2) that services were interrupted in 2024 and IRS was not reestablished (CF2). These scenarios were then used to estimate avertable cases occurring in 2024 (i.e. averted in CF1) and cases averted by the reintroduction of IRS (i.e. cases averted with respect to CF2). Confidence intervals of scenarios and avertable cases were calculated as the 95% credible interval based on a multivariate normal approximation of the posterior distribution of model coefficients [22]. No statistical adjustment for historical vector control coverage, or confounders other than precipitation was made, due to a lack of availability of these covariates at the temporal resolution required for inclusion in this model (e.g., LLIN use is available only on annual scale from the MIS).

Analysis of survey data followed a method which has been previously used on Bioko MIS data to analyze the impact of travel [24]. This consisted of fitting an interrupted time series logistic regression model to *Plasmodium falciparum* prevalence (*Pf*PR) data from 2019-2024, where the impact of the interruption of services was defined as the change observed in relation to the linear 2019-2023 trend. Results are reported as adjusted and unadjusted odds ratios (OR), where variables used in adjustment were recent history of travel (in the past 8 weeks), early house entry (before 7PM), SES quintile, and precipitation in the two months and four months prior to the MIS interview. A third model, referred to below as vector control as covariate (VCC) model, incorporated historical IRS coverage and LSM targeting at the PSU level, and LLIN use as reported in the MIS as covariates to explicitly quantify the effect of vector control interruptions in 2024 instead of a linear interruption term, in addition to other covariates. For this model, a counterfactual scenario for 2024 with no interruption was created, assuming IRS coverage and LSM targeting would remain unchanged from 2023 and setting LLIN use as the extrapolation of a linear trend from 2019-2023 for Malabo and Baney, and from 2021-2023 for Luba and Riaba (where a mass distribution campaign was conducted in 2021). Based on prior evidence from Bioko Island [17,25], IRS coverage was modeled as a categorical covariates with three levels (<30%, 30-49% and 50% or more) and due to difficulties is defining LSM coverage, LSM activities were captured through a binary variable (targeted for LSM vs not). To limit potential bias related to increased vector control targeting and coverage in high-transmission areas, 2018 PSU-level F*f*PR was included as an offset in the VCC model. Individuals with missing covariates were excluded from all analyses. In all models, the impact of interruptions, temporal trend, intercept, and, in the case of VCC, the effects of vector control, were fit via interaction terms to allow variation by district, given the likelihood of differing trends and effects. Throughout, survey sampling weights were used to ensure estimates are representative of Bioko Island.

## Results

### Changes in confirmed cases

Health facilities on Bioko reported a total of 15,121 confirmed malaria cases in 2024 (49.9 per 1,000 population), which represented an increase of 41% compared to the 2021-2023 average (10,761 per year, 37.4 per 1,000). Case incidence varied greatly by district and was an order of magnitude higher in Luba and Riaba than Malabo and Baney, both before (annual average 125.8 and 322.8 per 1,000 in Luba and Riaba versus 32.3 and 29.3 in Malabo and Baney) and after the interruption (105.2 and 311.5 versus 47.9 and 32.4, respectively). There was also a precipitous decline in cases in the final months of 2024, corresponding to the period after the reintroduction of IRS. Statistical modeling of counterfactual scenarios showed that the observed increase in incidence in 2024 was unexplained by previous trends, while the decline in reported cases from October 2024 onwards was associated with achieved IRS coverage **(Figure 1)**. Comparison of scenarios predicted that overall, 23.1% (95% CI 9.2-33.7%) of cases observed in 2024 were avertable, while the reestablishment of IRS in late 2024 was predicted to have averted an additional increase of 13.3% (95% CI 4.4-28.4%) **(Figure 2)**.

**Figure 1.**
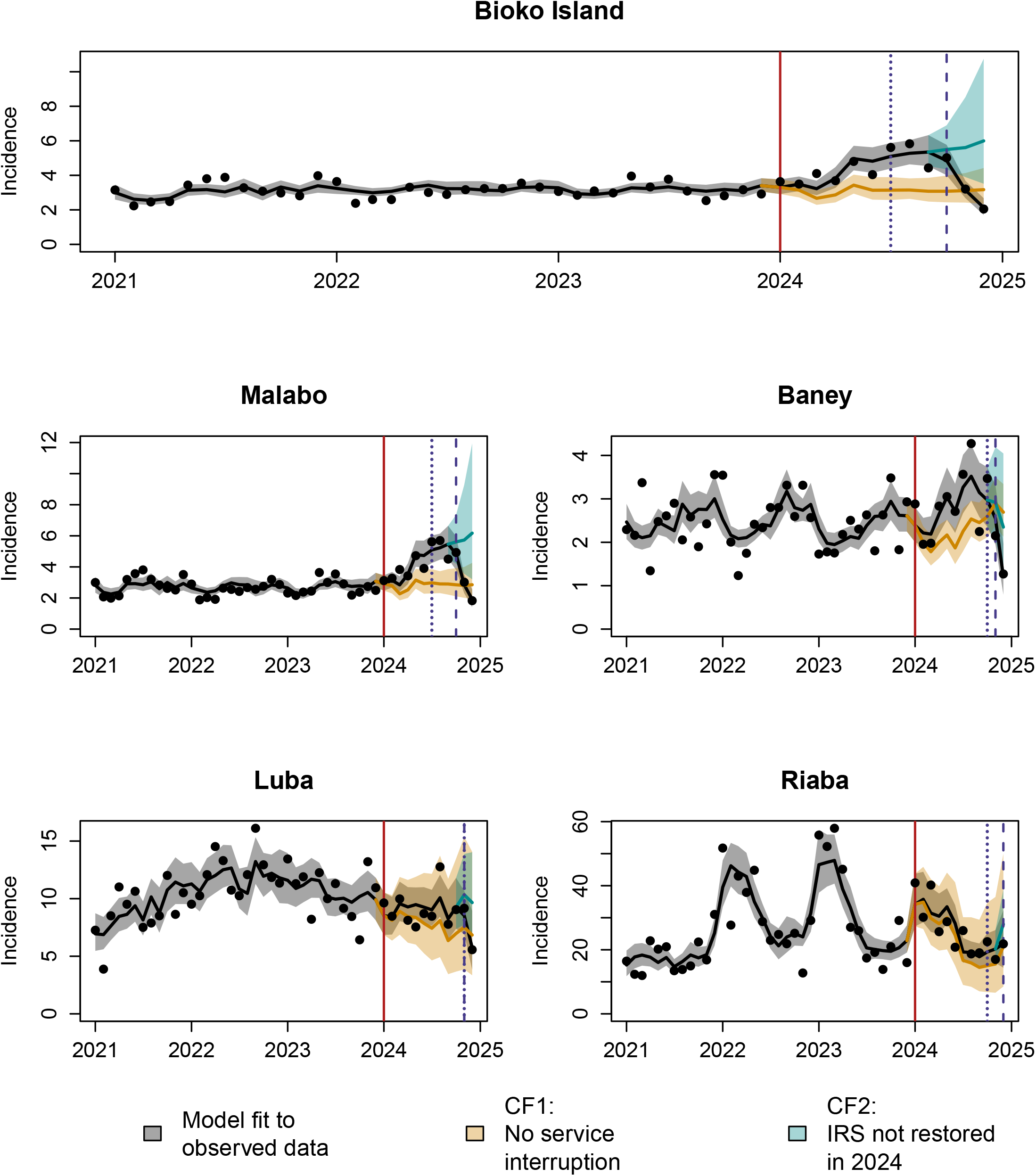
Monthly case incidence, fitted model and counterfactual scenarios. Points indicate the number of confirmed cases reported per 1,000 population, while the black, blue and orange lines show the fitted model, and counterfactual scenarios CF1 (no interruption of services in 2024) and CF2 (no reestablishment of IRS in 2024), respectively. 95% confidence intervals are shown as shaded bands around lines. Vertical lines show the timing of interruption of field activities (solid red line), and re-establishment of LLIN distribution (blue dotted line) and IRS (blue dashed line) by district.

**Figure 2:**
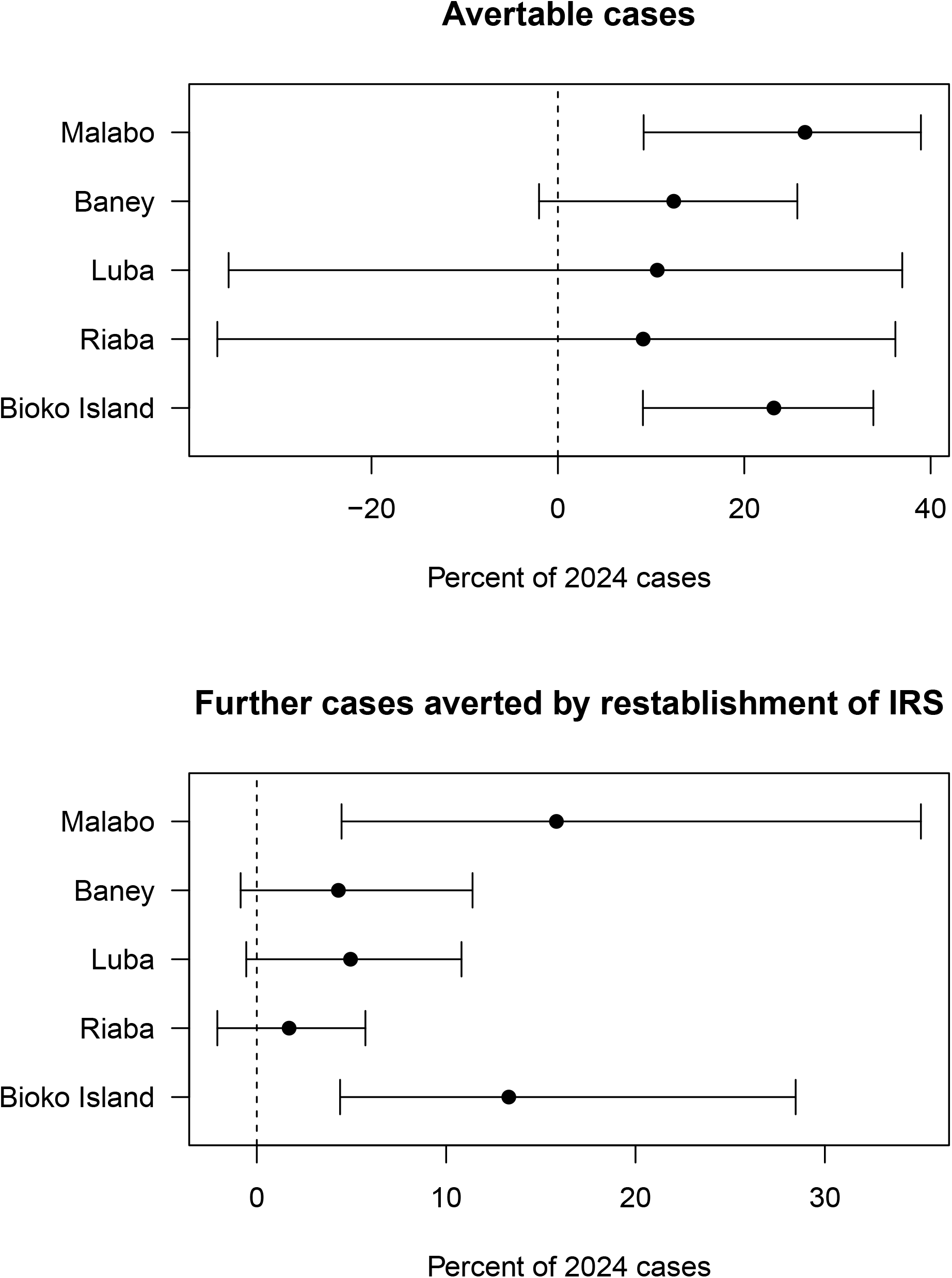
Estimated avertable cases and cases averted by reintroduction of IRS, by district. Avertable cases were calculated as the number of cases in 2024 which could have been averted if IRS had not been interrupted, and further cases averted were calculated as the number of cases which would have occurred during the study period had IRS not been reestablished in late 2024. In both cases, these numbers are standardized as the percentage of predicted 2024 cases. Points show mean and whiskers indicate the 95% confidence interval.

Notably, the impact of the interruption of malaria control and reintroduction of IRS varied across districts. Given its high population, estimates for Malabo were highly similar to those overall, with 26.5% of 2024 cases potentially avertable (95% CI 9.9-38.9%) and the reintroduction of IRS predicted to have averted a further increase of 15.8% (95% CI 4.5-34.9%). In other districts no statistically significant impact of the interruption or reestablishment of control activities was identified, although the mean estimates of avertable cases in Baney, Luba and Riaba districts (9.1-12.6% of cases reported in 2024) and cases averted by reestablishment of control in Luba and Baney (5.0% and 4.3% further increase in cases averted, respectively) were meaningful in magnitude **(Figure 2)**. The sensitivity analysis accounting for autocorrelation found similar results, with the notable exception that the re-establishment of IRS was found to be significantly associated with a decrease in incidence in Baney and Luba (although mean estimates were nearly unchanged compared to the main analysis, see Supplemental Information, Figure S4).

### Changes in prevalence

In the six years of MIS included in this analysis, 29,902 household surveys were conducted with 77,774 RDTs administered, and all covariates used in adjustment were available for the vast majority of these individuals **(Table 1)**. Comparison of the full study population and the analytical dataset revealed no significant shifts in parasite prevalence (PfPR), demographic distribution, or socioeconomic status. Given this high degree of similarity across all strata, data were considered missing completely at random (MCAR), and a complete-case analysis was performed. Overall, there was a three percentage-point increase in *Pf*PR from 12.9% in 2023 to 15.9% in 2024, primarily driven by an increase in Malabo district **(Figure 3)**. The overall increase was statistically significant and corresponded to an adjusted odds ratio of 1.20 above the 2019-2023 trend (95% CI 1.07-1.35, unadjusted OR 1.19,95% CI 1.06-1.33). However, at a district level the departure from previous trends was statistically significant only in Malabo, where the increase mirrored the overall results (adjusted OR 1.27, 95% CI 1.12-1.44) **(Figure 4)**. In Luba no significant increase beyond previous trends was identified (adjusted OR 1.16, 95% CI 0.75-1.79). Counterintuitively, Baney and Riaba registered lower *Pf*PR than expected from previous trends, although these were not statistically significant (adjusted OR 0.80 95% 0.58-1.11 and 0.80, 95% CI 0.55-1.35, respectively).

**Table 1.** Characteristics of MIS participants and covariates. Columns labelled ‘Analytical Dataset’ correspond to observations used in final (adjusted and unadjusted) analysis, with non-missing values for adjustment covariates. Note that individuals and households with unknown age or undefined SES quintile are excluded from the age and SES quintile breakdowns, respectively.

|  | All Data |  |  | Analytical Dataset |  |  |  |  |  |
| --- | --- | --- | --- | --- | --- | --- | --- | --- | --- |
|  | Households surveyed | Individuals tested | *PfPR* | Households surveyed | Individuals tested | *PfPR* | LLIN use | Early house entry | Travel history |
| **Year** |  |  |  |  |  |  |  |  |  |
| 2019 | 5,074 | 13,918 | 13.4 % | 4,402 | 12,639 | 13.1 % | 38.4 % | 28.4 % | 10.7 % |
| 2020 | 4,963 | 12,738 | 12.1 % | 3,971 | 11,236 | 11.7 % | 37.2 % | 22.2 % | 1.9 % |
| 2021 | 4,928 | 12,515 | 13.8 % | 4,064 | 11,018 | 13.5 % | 34.5 % | 27.9 % | 2.9 % |
| 2022 | 5,026 | 12,337 | 14.3 % | 4,141 | 10,769 | 13.9 % | 36.1 % | 32.3 % | 6.3 % |
| 2023 | 4,998 | 13,300 | 12.9 % | 4,170 | 11,490 | 12.4 % | 34.2 % | 32.8 % | 6.6 % |
| 2024 | 4,913 | 12,966 | 15.9 % | 4,105 | 11,381 | 15.2 % | 29.0 % | 29.3 % | 5.1 % |
| **District** |  |  |  |  |  |  |  |  |  |
| Malabo | 22,690 | 59,342 | 13.6 % | 18,556 | 52,189 | 13.2 % | 35.2 % | 28.0 % | 5.9 % |
| Baney | 3,709 | 10,260 | 12.9 % | 3,178 | 9,053 | 12.6 % | 31.3 % | 34.6 % | 5.2 % |
| Luba | 2,150 | 4,671 | 16.4 % | 1,904 | 4,202 | 16.0 % | 47.8 % | 23.1 % | 2.5 % |
| Riaba | 1,353 | 3,501 | 21.8 % | 1,215 | 3,089 | 21.5 % | 36.4 % | 25.8 % | 2.9 % |
| **SES quintile** |  |  |  |  |  |  |  |  |  |
| Lowest | 7,400 | 12,817 | 16.6 % | 5,961 | 11,274 | 16.3 % | 39.0 % | 21.6 % | 3.2 % |
| Second | 6,017 | 14,848 | 16.0 % | 5,053 | 13,115 | 15.5 % | 42.5 % | 25.8 % | 4.0 % |
| Middle | 5,495 | 15,537 | 14.6 % | 4,662 | 13,683 | 14.1 % | 39.9 % | 27.1 % | 4.8 % |
| Fourth | 5,544 | 16,839 | 13.0 % | 4,727 | 14,892 | 12.4 % | 36.7 % | 30.7 % | 5.8 % |
| Highest | 5,421 | 17,686 | 10.4 % | 4,450 | 15,569 | 10.2 % | 21.3 % | 34.5 % | 9.0 % |
| **Age** |  |  |  |  |  |  |  |  |  |
| <5 |  | 10,777 | 8.2 % |  | 9,643 | 8.0 % | 41.2 % | 44.1 % | 3.1 % |
| 5-14 |  | 23,214 | 17.1 % |  | 20,220 | 16.7 % | 35.6 % | 33.7 % | 3.0 % |
| 15+ |  | 43,342 | 13.3 % |  | 38,261 | 12.9 % | 32.8 % | 22.0 % | 7.8 % |
| **Overall** | **29,902** | **77,774** | **13.7 %** | **24,853** | **68,533** | **13.3 %** | **34.9 %** | **28.8 %** | **5.7 %** |

**Figure 3.**
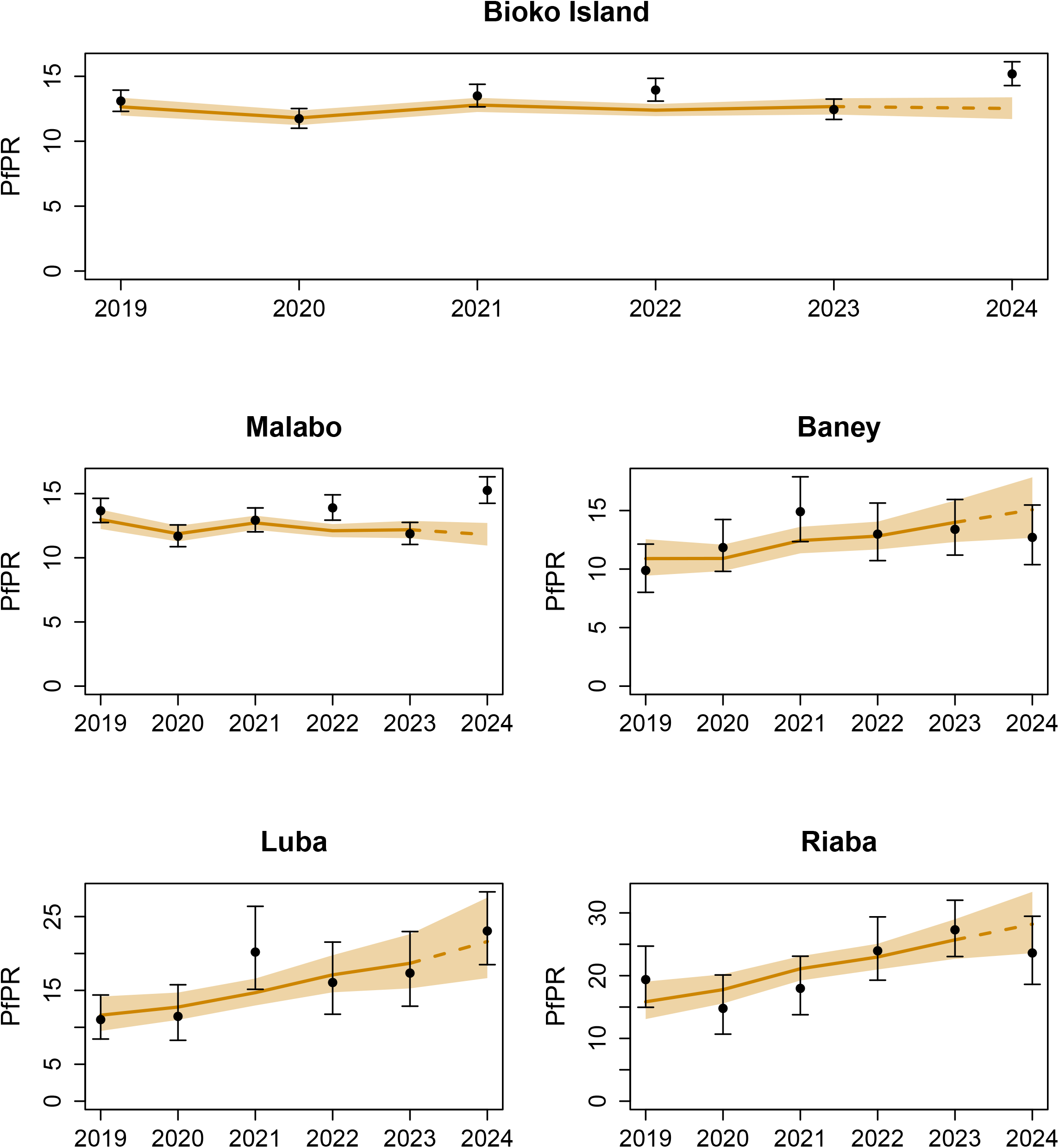
Estimated and observed *Pf*PR overall and by district from 2019-2024. Solid line and ribbon indicate adjusted model fit and 95% CI, and the dashed line corresponds the 2019-2023 trend extrapolated to 2024. Points and whiskers indicate observed *Pf*PR.

**Figure 4.**
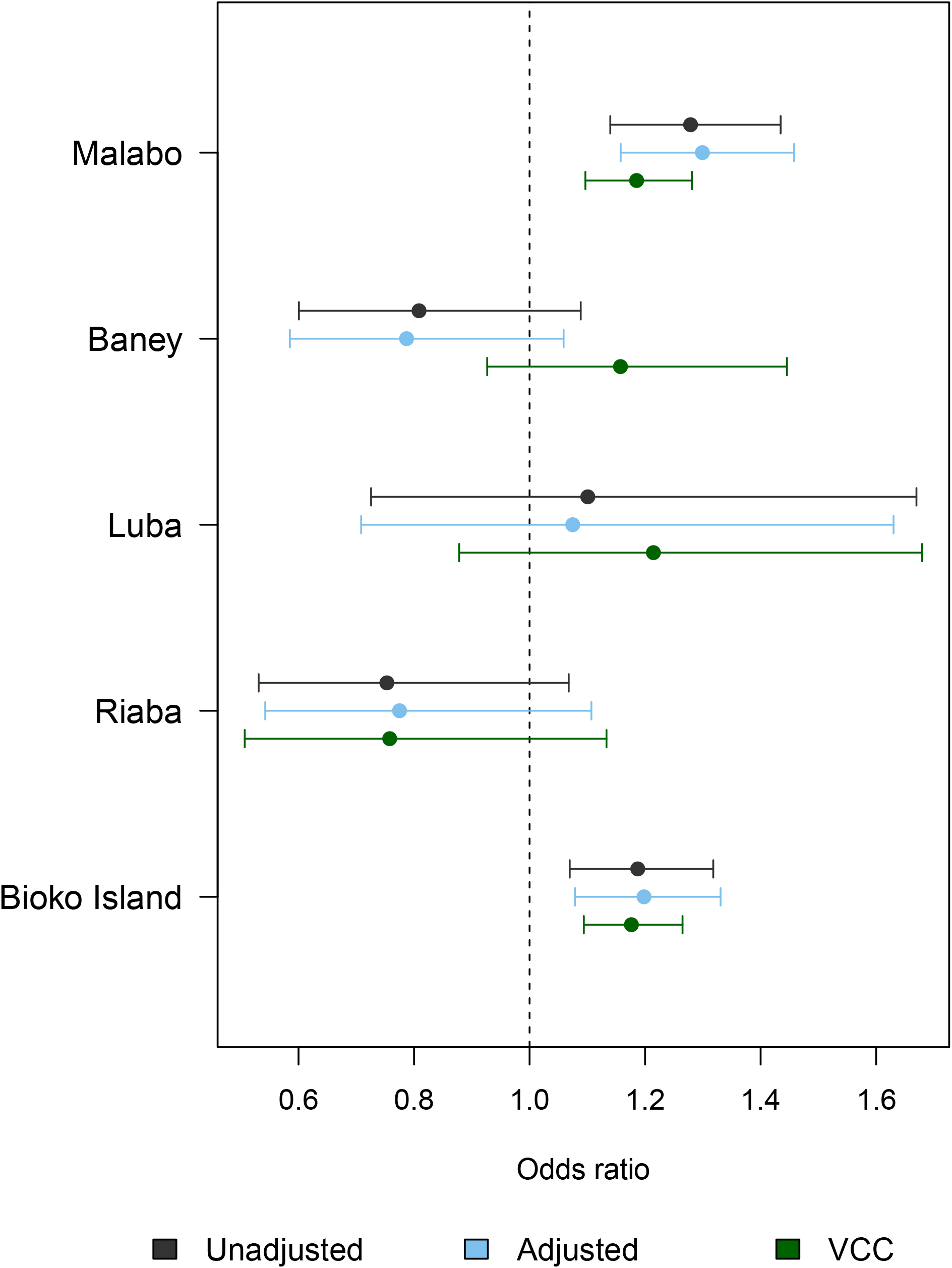
Odds ratios (OR) of P*f*PR attributed to the interruption of malaria control activities in 2024 according to unadjusted, adjusted and vector control as covariate (VCC) models, compared to 2019-2023 trend, by district and overall. Covariates used in adjustment were recent travel history, early house entry and SES quintile, while vector control as covariate model also included historical IRS coverage, LLIN use and LSM targeting. Points show estimates, whiskers indicate the 95% CI, and the vertical dotted line corresponds to no effect (OR=1).

As shown in **Table 2**, there was a significant change in achieved vector control coverage in 2019-2020 versus 2021-2023, corresponding to the expansion of IRS to nearly all areas of the island and implementation of LSM in Malabo and Baney districts. The VCC model found that IRS coverage in Malabo and Baney, and LLIN use in all districts except Luba was significantly associated with lower odds of infection, while associations with LSM targeting, LLIN use in Luba and IRS coverage in Luba and Riaba were not statistically significant. Estimates of the impact of the interruption in the VCC model were highly similar to the those of the other models for Bioko Island overall **(Figure 4**, OR 1.19, 95% 1.10-1.29), and in all districts except for Baney where the direction of effect was different, although still not statistically significant (OR 1.21, 95% CI 0.95-1.54).

**Table 2.** Vector control coverage achieved from 2019-2024, by district. Percentages indicate the roportion of the population living in PSUs with PSU-level IRS coverage at indicated levels, diich had any part of the PSU targeted for LSM by district and year, and who reported sleeping nder a LLIN the night prior to the MIS interview. Values in 2024 correspond to period up to eginning of MIS fieldwork, so IRS coverage is zero in all districts. Estimation conducted only on on-missing MIS records.

| Year | District | IRS coverage:<br><30% or not<br>targeted | IRS coverage:<br>30-50% | IRS coverage:<br>≥50% | Targeted for<br>LSM | LLIN use |
| --- | --- | --- | --- | --- | --- | --- |
| 2019 | Baney | 77.8% | 10.2% | 12.1% | 0% | 27.6% |
|  | Luba | 27.1% | 0% | 72.9% | 0% | 56.0% |
|  | Malabo | 69.4% | 16.6% | 14.0% | 0% | 39.6% |
|  | Riaba | 0% | 0% | 100.0% | 0% | 35.1% |
| 2020 | Baney | 63.2% | 18.7% | 18.1% | 0% | 35.1% |
|  | Luba | 52.7% | 0% | 47.3% | 0% | 48.7% |
|  | Malabo | 31.4% | 26.5% | 42.2% | 0% | 37.2% |
|  | Riaba | 0% | 0% | 100.0% | 0% | 41.2% |
| 2021 | Baney | 17.0% | 27.7% | 55.3% | 0% | 36.6% |
|  | Luba | 5.9% | 0% | 94.1% | 0% | 55.0% |
|  | Malabo | 4.5% | 18.6% | 77.0% | 5.8% | 33.1% |
|  | Riaba | 0% | 0% | 100.0% | 0% | 51.5% |
| 2022 | Baney | 18.6% | 35.3% | 46.1% | 64.6% | 33.6% |
|  | Luba | 3.8% | 0% | 96.2% | 0% | 45.1% |
|  | Malabo | 6.2% | 28.4% | 65.4% | 87.3% | 36.3% |
|  | Riaba | 0% | 0% | 100.0% | 0% | 37.5% |
| 2023 | Baney | 0% | 51.2% | 48.8% | 82.7% | 32.3% |
|  | Luba | 0% | 0% | 100.0% | 0% | 36.8% |
|  | Malabo | 21.3% | 40.5% | 38.3% | 88.9% | 34.5% |
|  | Riaba | 0% | 0% | 100.0% | 0% | 31.6% |
| 2024 | Baney | 100.0% | 0.0% | 0.0% | 0.0% | 22.6% |
|  | Luba | 100.0% | 0.0% | 0.0% | 0.0% | 38.8% |
|  | Malabo | 100.0% | 0.0% | 0.0% | 0.0% | 29.8% |
|  | Riaba | 100.0% | 0.0% | 0.0% | 0.0% | 25.2% |

## Discussion

The results reported here demonstrate the speed with which gains from malaria control can be lost, but also that reintroduction of control can regain lost ground just as quickly. In a period of only one year without IRS on Bioko, cases rose by nearly 50%, while prevalence returned to levels last seen around 10 years ago [26]. Unsurprisingly, these changes were significant departures from the pre-interruption trends, adding new evidence that resurgence tends to occur after interrupting control operations [5]. Even on Bioko, this is not the first documented impact of funding interruptions. In 2019, an outbreak was registered in Riaba, at least partially caused by a delay in IRS implementation due to a lapse in funding [27]. However, the 2024 funding disruption was substantially longer and the evidence from this study shows it had more widespread effects, including in the urban portion of the island (principally, in Malabo district). More importantly, the data presented here demonstrate that while resurgences are rapid, they can also be quickly contained by the reintroduction of control activities. Despite substantial resurgence in reported cases, by the end of 2024 the reestablishment of control helped to reduce monthly case counts to a level comparable to what would have been expected with no interruption.

Interestingly, analyses of cases and prevalence data did not entirely agree. Both analyses found a higher than otherwise expected burden in 2024 overall and for Malabo district, and no statistically significant change in Baney, Luba or Riaba. However, meaningful (although not statistically significant) decreases in prevalence compared to previous trends were found in Riaba and possibly Baney, while analysis of confirmed cases showed non-statistically significant increases in 2024 incidence in these districts. In the case of Riaba, this inconsistency was likely caused by a decreasing interannual trend starting from late 2022 to early 2023, which could be identified in case data due to their higher temporal resolution, but not in MIS data since only a single data point was available between the beginning of 2023 and the interruption of vector control. In Baney, there was uncertainty about even the direction of effects found by prevalence models, with the VCC mean estimate suggesting an increase while the adjusted model suggested a decrease. This discrepancy was likely caused by the fact that vector control coverage pre-interruption was found to be protective of malaria infection, but prevalence remained nearly unchanged after the interruption. Qualitatively differing findings such as these are indicative of genuine uncertainty about underlying effects and suggest that there may be unaccounted factors playing an important role, warranting further investigation. More broadly, the higher uncertainty in estimates of the impact of the interruption in Luba and Riaba may be related to their much higher level of transmission and small population, resulting in higher variance. It is also possible that due to their high transmission level, even a substantial real change in transmission intensity could result in limited changes in all-age incidence, given the saturating relationship between transmission intensity and incidence, especially among older children and adults, identified by mathematical modeling [28].

It is important to emphasize that the increase in burden reported here is not reflective of a full interruption of malaria control activities. Since the disruption lasted only one year, the impact on LLIN use was limited **(Tables 1-2)**. Had LLIN use been higher, the effects of this interruption may have been lesser (and indeed, the relatively higher use of LLINs in Luba may have limited the impact of the interruption), although high or even moderate LLIN use on Bioko has shown to be difficult to sustain [16]. Temporary agreements with funders also limited the impact that the disruption had on routine case management, for example avoiding stockouts of diagnostic tools and antimalarial treatments. Thus, the effects observed are primarily due to an interruption of IRS and urban LSM and would have almost certainly been larger for a longer lasting or more complete interruption. A similarly rapid resurgence of cases following a cessation of IRS was observed in Uganda, although the interruption was longer and the effect was larger than that observed here [6]. The impact of such disruptions on prevalence is less well-studied, mostly due to the difficulty of collecting prevalence data during funding lapses. However, the overall increase in *PfPR* reported here is broadly consistent with the effect size of IRS estimated from a historical analysis of Bioko Island MIS data [25], further supporting the interpretation that these changes are caused in large part by the interruption to IRS. This is corroborated by the fact that in the VCC model, the impact of the interruption was driven by the effect size of IRS, since changes to LLIN use were limited and LSM targeting was not found to have a statistically significant association with prevalence. More broadly, the effect sizes observed here may be particular to the context of Bioko, with its long history of intense control (principally, IRS).

The main weakness of this study is the assumption that, when extrapolated to 2024, observed pre-interruption trends provide a reasonable estimate of what would have occurred had activities not been interrupted. In particular, the opening of a new public hospital in 2024 is potentially problematic for the attribution of changes in reported cases to control interruptions. However, the rapid decline in cases after the reestablishment of IRS in late 2024, including those reported from the General Hospital of Sampaka, suggests that increases in confirmed cases are more likely to have been caused by the lack of vector control than changes in the health system. Similarly, the use of linear temporal trends for MIS analyses may have obscured important pre-interruption variation, although this does not appear to be a major issue on visual inspection, with the possible exclusion of Riaba **(Figure 3)**. Additionally, the small populations of Luba and Riaba limit the interpretation of results for these districts. Despite these weaknesses, the overall consistency of results of case and prevalence analyses, and especially the impact on cases seen after the reestablishment of IRS in late 2024 provide strong evidence that the increase in burden observed was caused at least in part by the interruption of vector control.

## Conclusion

This study provides a clear and well-documented case of the impacts that interruptions to malaria control and reintroductions of control can have. Substantial increases in confirmed malaria cases and malaria prevalence were observed after less than one year with limited control. While this resurgence was rapid, it was also quickly contained and reversed by the reintroduction of control activities. These are important considerations for funders when taking decisions on reducing or re-allocating malaria control funding, as well as when responding to subsequent resurgences.

## Acknowledgements

We thank the staff of public health facilities on Bioko Island, data digitizers, and fieldworkers and participants of the annual Malaria Indicator Survey (MIS) from 2019-2024, without whom this analysis would not have been possible.

## Ethics approval

The protocol for the annual Malaria Indicator Survey was approved by the Technical and Ethics Committee of the Ministry of Health and Social Welfare of Equatorial Guinea and the Ethics Committee of the London School of Hygiene and Tropical Medicine. All adult survey participants provided written informed consent for themselves and, where relevant, on behalf of children under 18 years of age in their household.

## Data availability

All HMIS, MIS, and vector control data and code used in this study are available at the GitHub repository https://github.com/galickda/bioko_interruption_2024. Precipitation data are available on request for non-commercial use from https://www.gloh2o.org/.

## Author contributions

DSG, DEBH, CAG and GAG conceived and designed the study. NBC and JS maintained and oversaw data collection in routine reporting systems (DHIS2). DSG and TAOM designed and oversaw data collection in the MIS. DSG and DEBH analyzed data and prepared figures. DLS, CAG and GAG provided technical and scientific guidance on study design, implementation and data analysis. MRR and WPP supervised implementation in Equatorial Guinea.

## References

1. World Health Organization. World malaria report 2024: addressing inequity in the global malaria response. Geneva: World Health Organization; 2024. Report No.

2. Bhatt S, Weiss DJ, Cameron E, Bisanzio D, Mappin B, Dalrymple U, et al. The effect of malaria control on Plasmodium falciparum in Africa between 2000 and 2015. Nature. 2015 Oct;526(7572):207–11. doi:10.1038/nature15535

3. Feachem RGA, Chen I, Akbari O, Bertozzi-Villa A, Bhatt S, Binka F, et al. Malaria eradication within a generation: ambitious, achievable, and necessary. The Lancet. 2019 Sep;394(10203):1056–112. doi:10.1016/S0140-6736(19)31139-0

4. Lawal L, Buhari AO, Jaji TA, Äiatare AS, Adeyemo AO, Olumoh AO, et al. Lingering challenges in malaria elimination efforts in sub-Saharan Africa: Insights and potential solutions. Health Sci Rep. 2024;7(6):e2122. doi:10.1002/hsr2.2122

5. Cohen JM, Smith DL, Cotter C, Ward A, Yamey G, Sabot OJ, et al. Malaria resurgence: a systematic review and assessment of its causes. Malar J. 2012 Apr 24;11:122. doi:10.1186/1475-2875-11-122 PubMed PMID: 22531245; PubMed Central PMCID: PMC3458906.

6. Namuganga JF, Epstein A, Nankabirwa JI, Mpimbaza A, Kiggundu M, Sserwanga A, et al. The impact of stopping and starting indoor residual spraying on malaria burden in Uganda. Nat Commun. 2021 May 11;12(1):2635. doi:10.1038/s41467-021-22896-5

7. Janko MM, Recalde-Coronel GC, Damasceno CP, Salmón-Mulanovich G, Barbieri AF, Lescano AG, et al. The impact of sustained malaria control in the Loreto region of Peru: a retrospective, observational, spatially-varying interrupted time series analysis of the PAMAFRO program. Lancet Reg Health - Am. 2023 Apr 1;20. doi:10.1016/j.lana.2023.100477

8. Dzianach PA, Rumisha SF, Lubinda J, Saddler A, van den Berg M, Gelaw YA, et al. Evaluating COVID-19-Related Disruptions to Effective Malaria Case Management in 2020-2021 and Its Potential Effects on Malaria Burden in Sub-Saharan Africa. Trop Med Infect Dis. 2023 Apr;8(4):4. doi:10.3390/tropicalmed8040216

9. Winskill P, Slater HC, Griffin JT, Ghani AC, Walker PGT. The US President’s Malaria Initiative, Plasmodium falciparum transmission and mortality: A modelling study. PLOS Med. 2017 Nov 21;14(11):e1002448. doi:10.1371/journal.pmed.1002448

10. Yukich JO, Chitnis N. Modelling the implications of stopping vector control for malaria control and elimination. Malar J. 2017 Oct 13;16(1):411. doi:10.1186/s12936-017-2051-1

11. Sherrard-Smith E, Hogan AB, Hamlet A, Watson OJ, Whittaker C, Winskill P, et al. The potential public health consequences of COVID-19 on malaria in Africa. Nat Med. 2020 Sep;26(9):1411–6. doi:10.1038/s41591-020-1025-y

12. García GA, Hergott DEB, Phiri WP, Perry M, Smith J, Osa Nfumu JO, et al. Mapping and enumerating houses and households to support malaria control interventions on Bioko Island. Malar J. 2019 Aug 22; 18(1): 1. doi:10.1186/s12936-019-2920-x

13. Fries B, Guerra CA, García GA, Wu SL, Smith JM, Oyono JNM, et al. Measuring the accuracy of gridded human population density surfaces: A case study in Bioko Island, Equatorial Guinea. PLOS ONE. 2021 Sep 1;16(9):9. doi:10.1371/journal.pone.0248646

14. Kleinschmidt I, Schwabe C, Benavente L, Torrez M, Ridl FC, Segura JL, et al. Marked Increase in Child Survival after Four Years of Intensive Malaria Control. Am J Trop Med Hyg. 2009 Jun;80(6):6. PubMed PMID: 19478243; PubMed Central PMCID: PMC3748782.

15. Overgaard HJ, Reddy VP, Abaga S, Matias A, Reddy MR, Kulkarni V, et al. Malaria transmission after five years of vector control on Bioko Island, Equatorial Guinea. Parasit Vectors. 2012 Nov 12;5(1):1. doi:10.1186/1756-3305-5-253

16. García GA, Galick DS, Smith JM, Iyanga MM, Rivas MR, Eyono JNM, et al. The challenge of improving long-lasting insecticidal nets coverage on Bioko Island: using data to adapt distribution strategies. Malar J. 2024 Oct 29;23(1):324. doi:10.1186/sl2936-024-05139-y

17. García GA, Hergott DEB, Galick DS, Donfack OT, Motobe Vaz L, Nze Nchama LO, et al. Testing indoor residual spraying coverage targets for malaria control, Bioko, Equatorial Guinea. Bull World Health Organ. 2025 Jun 1;103(6):392–402. doi:10.2471/BLT.24.292505 PubMed PMID: 40511394; PubMed Central PMCID: PMC12161160.

18. Cook J, Hergott D, Phiri W, Rivas MR, Bradley J, Segura L, et al. Trends in parasite prevalence following 13 years of malaria interventions on Bioko island, Equatorial Guinea: 2004-2016. Malar J. 2018 Feb 5;17(1):1. doi:10.1186/s12936-018-2213-9

19. Roll Back Malaria. Malaria Indicator Survey Toolkit [Internet]. Available from: https://www.malariasurveys.org/toolkit.cfm

20. Citron DT, Guerra CA, García GA, Wu SL, Battle KE, Gibson HS, et al. Quantifying malaria acquired during travel and its role in malaria elimination on Bioko Island. Malar J. 2021 Aug 30;20(1):1. doi:10.1186/s12936-021-03893-x

21. Wang X, Alharbi RS, Baez-Villanueva OM, Miralles DG, Ma J, Xu S, et al. MSWEP V3: Machine Learning-Powered Global Precipitation Estimates at 0.1° Hourly Resolution (1979-Present) [Internet]. arXiv; 2026 [cited 2026 Mar 6]. Available from: http://arxiv.org/abs/2602.01436 doi:10.48550/arXiv.2602.01436

22. Wood SN. Generalized Additive Models: An Introduction with R. 2nd ed. Chapman and Hall/CRC; 2017.

23. García GA, Atkinson B, Donfack OT, Hilton ER, Smith JM, Eyono JNM, et al. Real-time, spatial decision support to optimize malaria vector control: The case of indoor residual spraying on Bioko Island, Equatorial Guinea. PLOS Digit Health. 2022 May 12;1(5):e0000025. doi:10.1371/journal.pdig.0000025

24. Hergott DEB. Impact of six-month COVID-19 travel moratorium on Plasmodium falciparum prevalence on Bioko Island, Equatorial Guinea. Nat Commun. 2024.

25. Galick DS, Vaz LM, Ondo L, Iyanga MM, Bikie FEE, Avue RMN, et al. Reconsidering indoor residual spraying coverage targets: A retrospective analysis of high-resolution programmatic malaria control data. Proc Natl Acad Sci. 2025 Apr 22;122(16):e2421531122. doi:10.1073/pnas.2421531122

26. National Malaria Control Program of Equatorial Guinea, MCD Global Health. Bioko Island Malaria Indicator Survey 2024. 2025. Report No.

27. Guerra CA, Fuseini G, Donfack OT, Smith JM, Ondo Mifumu TA, Akadiri G, et al. Malaria outbreak in Riaba district, Bioko Island: lessons learned. Malar J. 2020 Aug 3;19(1):277. doi:10.1186/s12936-020-03347-w

28. Cameron E, Battle KE, Bhatt S, Weiss DJ, Bisanzio D, Mappin B, et al. Defining the relationship between infection prevalence and clinical incidence of Plasmodium falciparum malaria. Nat Commun. 2015 Sep 8;6(1):8170. doi:10.1038/ncomms9170

