## Supplementary Information for "Impact of a one-year interruption to vector control on Bioko Island, Equatorial Guinea"

**Sensitivity analysis of incidence models**

To assess the potential importance of autocorrelation in analyses of case incidence, auto-correlation function (ACF) and partial autocorrelation function (PACF) plots for residuals of the fitted model presented in the main text by district were examined (**Figure S1**). Since they show only minor possible residual autocorrelation, the original model was retained. However, additional analyses accounting for autocorrelation were also performed.

First, AR1 coefficients for each district as computed for the ACF plots were examined. Since they differed in both magnitude and sign, further analyses were performed at the district level. For each district, a model equivalent in structure to the main text model was constructed, incorporating a AR1 correlation structure with known correlation coefficient ($\rho$). Since the original model structure incorporated interactions for all terms (i.e. did not include any global effects other than the intercept), this is effectively no change to model structure other than the AR1 correlation term. To identify $\rho$ for each district, an optimization was run on the AIC of the fitted model. After identifying $\rho$, the residuals were once again examined for potential residual autocorrelation (**Figure S2**).

Finally, model estimates were computed in the same manner as in the main text, and results compared (**Figures S3-S4**). As noted in the main text, the only significant departure from models presented in the main text is the significance of the estimated increase in cases associated with the interruption of vector control in Baney and Luba districts. While significance differed, the magnitude of effects, including for Baney and Luba, was highly similar.

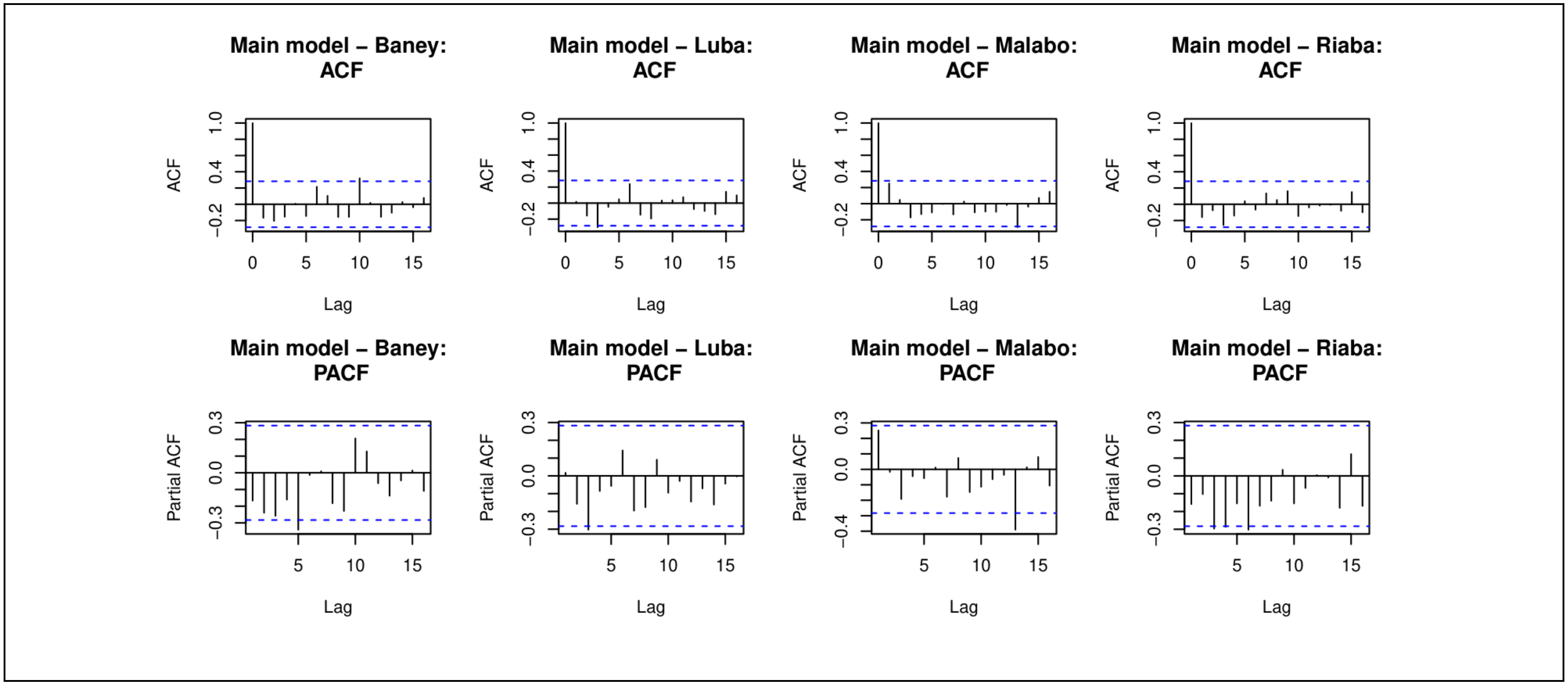


**Figure S1**: Autocorrelation and Partial autocorrelation plots (ACF, PACF, respectively) of the residuals from the incidence model presented in the main text, by district.

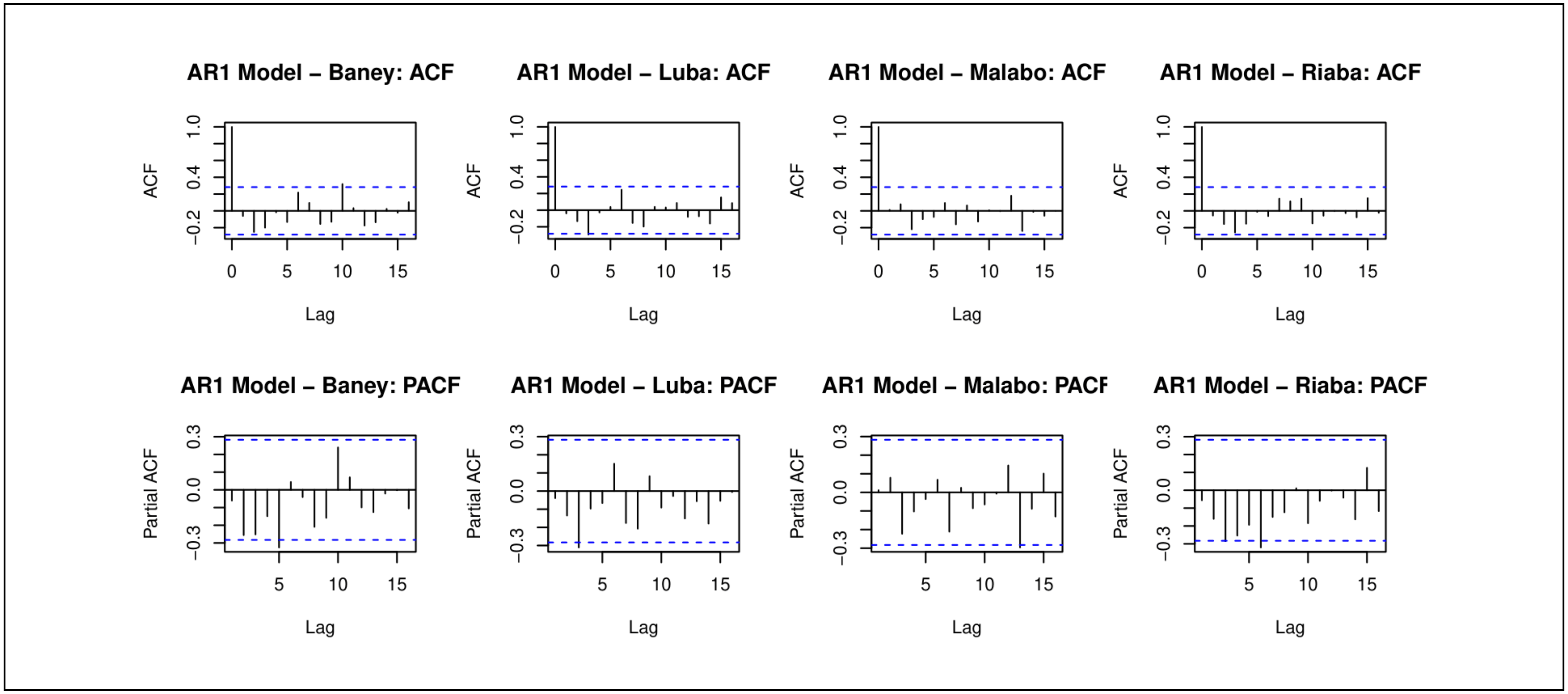


**Figure S2**: Autocorrelation and Partial autocorrelation plots (ACF, PACF, respectively) of the residuals from incidence models incorporating AR1 correlation structures, as described in this Supplemental Material.

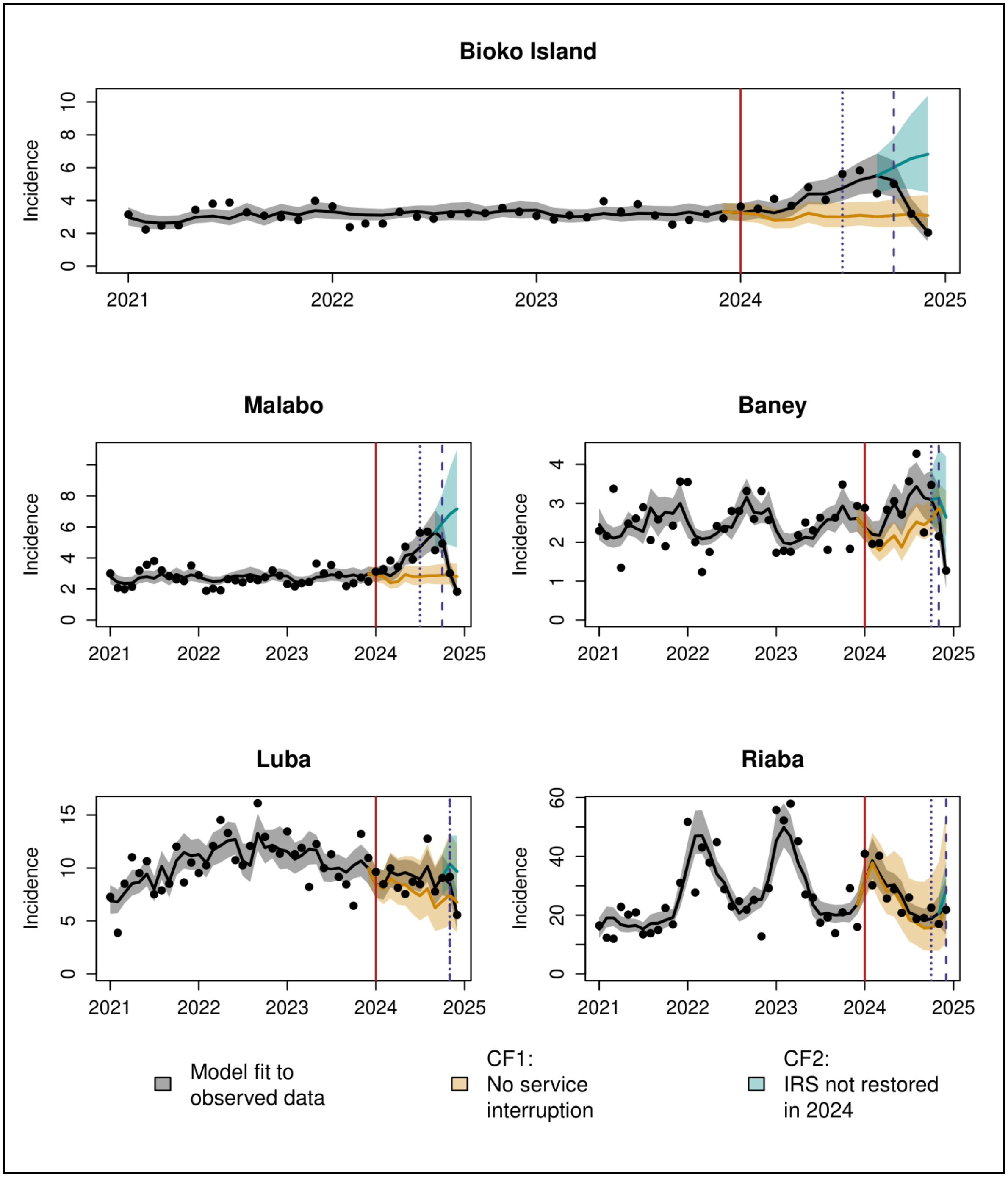


**Figure S3**: Monthly case incidence, fitted model and counterfactual scenarios, based on model fitted with with AR1 correlation structure (comparable to main text Figure 1). Points indicate the number of confirmed cases reported per 1,000 population, while the black, blue and orange lines show the fitted model, and counterfactual scenarios CF1 (no interruption of services in 2024) and CF2 (no reestablishment of IRS in 2024), respectively. 95% confidence intervals are shown as shaded bands around lines. Vertical lines show the timing of interruption of field activities (solid red line), and re-establishment of LLIN distribution (blue dotted line) and IRS (blue dashed line) by district.

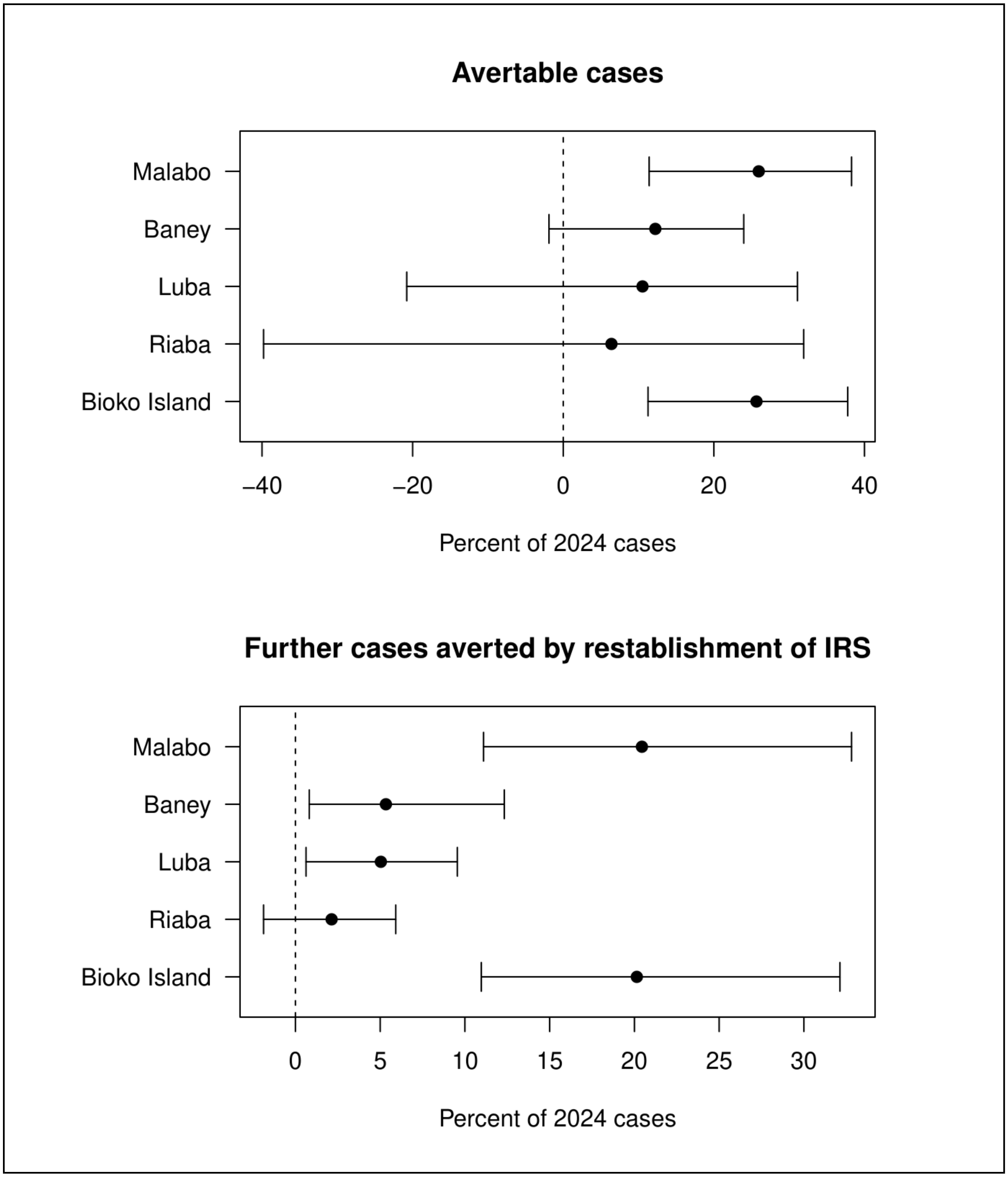


**Figure S4**: Estimated avertable cases and cases averted by reintroduction of IRS, by district, based on model fitted with AR1 correlation structure (comparable to main text Figure 2). Avertable cases were calculated as the number of cases in 2024 which could have been averted if IRS had not been interrupted, and further cases averted were calculated as the number of cases which would have occurred during the study period had IRS not been reestablished in late 2024. In both cases, these numbers are standardized as the percentage of predicted 2024 cases. Points show mean and whiskers indicate the 95% confidence interval.
